# The Activin/Follistatin-axis is severely deregulated in COVID-19 and independently associated with in-hospital mortality

**DOI:** 10.1101/2020.09.05.20184655

**Authors:** Evgenia Synolaki, Vasileios Papadopoulos, Georgios Divolis, Olga Tsahouridou, Efstratios Gavriilidis, Georgia Loli, Ariana Gavriil, Christina Tsigalou, Nikolaos R. Tziolos, Eleni Sertaridou, Bhanu Kalra, Ajay Kumar, Petros Rafailidis, Arja Pasternack, Dimitrios T. Boumpas, Georgios Germanidis, Olli Ritvos, Simeon Metallidis, Panagiotis Skendros, Paschalis Sideras

**Affiliations:** Biomedical Research Foundation Academy of Athens, Center for Clinical, Experimental Surgery & Translational Research, Athens, Greece; First Department of Internal Medicine, University Hospital of Alexandroupolis, Democritus University of Thrace, Alexandroupolis, Greece; First Department of Internal Medicine, AHEPA University Hospital, Aristotle University of Thessaloniki, Thessaloniki, Greece; Laboratory of Microbiology, University Hospital of Alexandroupolis, Democritus University of Thrace, Alexandroupolis, Greece; Fourth Department of Internal Medicine, Attikon University Hospital, National and Kapodistrian University of Athens Medical School, Athens, Greece; Intensive Care Unit, University Hospital of Alexandroupolis, Alexandroupolis, Greece; AnshLabs, Webster, Texas, USA; Second Department of Internal Medicine, University Hospital of Alexandroupolis, Democritus University of Thrace, Alexandroupolis, Greece; Department of Physiology, Faculty of Medicine, University of Helsinki, Helsinki, Finland; Laboratory of Molecular Hematology, Democritus University of Thrace, Alexandroupolis, Greece

**Keywords:** COVID-19, SARS-CoV-2, Activin, Follistatin, outcome

## Abstract

**Background:** Activins are members of the TGFβ-superfamily implicated in the pathogenesis of several immuno-inflammatory disorders. Based on our previous studies demonstrating that over-expression of Activin-A in murine lung causes pathology sharing key features of COVID-19, we hypothesized that Activins and their natural inhibitor Follistatin might be particularly relevant to COVID-19 pathophysiology.

**Methods:** Activin-A, Activin-B and Follistatin levels were retrospectively analyzed in 574 serum samples from 263 COVID-19 patients hospitalized in three independent centers, and compared with common demographic, clinical and laboratory parameters. Optimal-scaling with ridge-regression was used to screen variables and establish a prediction model.

**Result:** The Activin/Follistatin-axis was significantly deregulated during the course of COVID-19, correlated with severity and independently associated with mortality. FACT-CLINYCoD, a novel disease scoring system, adding one point for each of Follistatin>6235pg/ml, Activin-A>591pg/ml, Activin-B>249pg/ml, CRP>10.3mg/dL, LDH>427U/L, Intensive Care Unit (ICU) admission, Neutrophil/Lymphocyte-Ratio>5.6, Years of Age>61, Comorbidities>1 and D-dimers>1097ng/ml, efficiently predicted fatal outcome in an initial cohort (AUC: 0.951±0.032, p<10^−6^). Two independent cohorts that were used for validation indicated comparable AUC (0.958, p=0.880 and 0.924, p=0.256, respectively).

**Conclusions:** This study unravels strong link between Activin/Follistatin-axis and COVID-19 mortality and introduces FACT-CLINYCoD, a novel pathophysiology-based tool that allows dynamic prediction of disease outcome, supporting clinical decision making.

## Introduction

COVID-19 pandemic constitutes presently the most prominent public-health issue worldwide. Thus far, severe acute respiratory syndrome coronavirus 2 (SARS-CoV-2) is responsible for ∼38 million COVID-19 cases and more than 1 million deaths [1]. The initial phase of COVID-19 relates to viral pathogenic effects and is characterized by mild “flu-like” symptoms [2]. However, one week after onset-of-symptoms, ∼20% of patients develop moderate-to severe-pneumonia often complicated by a cytokine release syndrome (CRS)-like hyper-inflammatory reaction that can lead to ARDS and multi-organ failure, thereby increasing substantially in-hospital mortality [2–5]. Various comorbidities and aging negatively affect COVID-19 outcome [3,6]. Accumulating evidence indicate that excessive activation of innate-immunity, deregulated neutrophils and thrombotic microangiopathy characterize the maladaptive host-response that drives COVID-19 pathophysiology [7–11]. These pathomechanisms lead to rapid progression of hypoxemic respiratory failure and protean clinical manifestations that involve almost every organ [12,13]. Monitoring hospitalized COVID-19 patients, in the midst of multiple continuously changing parameters is challenging [2,3,5,13]. Therefore, development of novel therapeutic strategies and bed-to-bench tools permitting day-to-day prediction of patient outcome is of utmost importance for such dynamically evolving and clinically heterogeneous disease [12,14–16].

Activin-A and Activin-B are members of the Transforming Growth Factor-β (TGF-β)-superfamily implicated in the regulation of numerous aspects of inflammation and/or tissue remodeling [17–19]. Follistatin, a physiological Activin inhibitor, binds to them, induces endocytosis and proteolytic degradation and modulates their bioavailability [20]. Activins and Follistatin, are synthesized continuously in healthy tissues [21], however, in immuno-inflammatory conditions, epithelial, endothelial, interstitial stroma cells and immune cells secrete higher levels that can be detected in serum as biomarkers of local or systemic stress [17–19].

We have previously described over-expression of Activin-A and Follistatin in bronchoalveolar lavage (BAL) of ARDS patients [18,22]. Moreover, we showed that ectopic expression of Activin-A in murine lungs causes ARDS-like pathology [22], which shares cardinal features of COVID-19 pathophysiology. These include, mobilization of neutrophils in the lung, alveolar epithelial and endothelial cell-loss, systemic hyper-coagulant state associated with increased Tissue-Factor mRNA levels and a cytokine-storm like response characterized by high levels of IL-6 and TNFα [5,7,8,18,23].

In view of these findings we hypothesized that the Activin/Follistatin-axis (A/F-axis) might be particularly relevant to COVID-19. To validate this hypothesis, we analyzed sera from COVID-19 patients and found that Activin-A, Activin-B and Follistatin were significantly upregulated during the crucial period when patients tend to deteriorate. Of note, A/F-axis components were independently associated with disease severity and in-hospital mortality. Based on that, we developed a scoring-system for prediction and monitoring of COVID-19 outcome in real-life using Activins, Follistatin and common clinical/laboratory parameters.

## Materials and methods

### Study design

This is a retrospective study with a single endpoint, the final outcome (survival or death). Three national reference hospitals from distant regions of Greece participated in the study (Supplementary Figure 1A). An initial cohort of 117 consecutive COVID-19 patients hospitalized at University Hospital, Alexandroupolis and “AHEPA” Hospital, Thessaloniki from March 10, 2020 and had an outcome until July 7, 2020 was endorsed and 314 randomly acquired samples were analyzed (Table 1 and Supplementary Figure 1B). Two independent validation cohorts belonging to the “second COVID-19 wave” between end of July and early October 2020 were introduced: The first included 28 consecutive COVID-19 patients derived from a distinct hospital (“Attikon” Hospital, Athens) contributing 35 samples and the second included 118 consecutive patients derived from Alexandroupolis and “AHEPA” hospitals contributing 225 samples (Supplementary Table 1). Activin-A, Activin-B, Follistatin, and standard-of-care (SOC) laboratory parameters including absolute-neutrophil-count (ANC), absolute-lymphocyte-count (ALC), neutrophil/lymphocyte-ratio (NLR), C-reactive protein (CRP), lactate dehydrogenase (LDH), ferritin and D-dimers, were analyzed. The study conformed to the TRIPOD statement [24] and is aligned with the Helsinki declaration. The study protocol design was approved by the Local Scientific and Ethics Committees and Institutional Review Boards of the University Hospital of Alexandroupolis (Ref. No. 803/23-09-2019 and Ref. No. 87/08-04-2020), AHEPA University Hospital of Thessaloniki (Ref. No. 1789/2020) and ATTIKON University Hospital of Athens (Ref. No. 487/3-9-2020). Patients’ records were anonymized and de-identified prior to analysis so confidentiality and anonymity were assured.

**Table 1.** Characteristics of patients (n=117) and samples (n=314).

| Parameter | patients (n=117) | samples (n=314) | P |
| --- | --- | --- | --- |
| **Age (years)** |  |  |  |
| Mean ($\pm 95\%$ CI) | 61.3±2.9 | 61.6±1.7 | 0.863 |
| **Sex** |  |  |  |
| Male | 63 (53.8) | 161 (51.3) | 0.635 |
| Female | 54 (46.2) | 153 (48.7) |  |
| **Number of comorbidities** |  |  |  |
| 0 | 28 (24.0) | 69 (22.0) | 0.813 |
| 1 | 33 (28.1) | 79 (25.2) |  |
| 2 | 26 (22.7) | 72 (22.9) |  |
| 3 | 21 (17.9) | 60 (19.1) |  |
| 4 | 6 (5.1) | 28 (8.9) |  |
| 5 | 3 (2.6) | 6 (1.9) |  |
| **Disease status (DS) at sampling** |  |  |  |
| DS1 | 39 (33.4) | 95 (30.2) | 0.363 |
| DS2 | 27 (23.1) | 74 (23.6) |  |
| DS3 | 32 (27.4) | 71 (22.6) |  |
| DS4 | 19 (16.2) | 74 (23.6) |  |
| **Treatment at sampling** |  |  |  |
| Hydroxychloroquine | 58 (49.6) | 98 (31.2) | 0.358 |
| Azithromycin | 69 (59.0) | 125 (39.8) |  |
| Antibiotics (other) | 103 (88.0) | 257 (81.8) |  |
| Lopinavir/Ritonavir | 57 (48.7) | 112 (35.7) |  |
| Remdesivir | 4 (3.4) | 10 (3.2) |  |
| Tocilizumab | 3 (2.6) | 5 (1.6) |  |
| Anakinra | 12 (10.3) | 31 (9.9) |  |
| LMWH | 91 (77.8) | 254 (80.9) |  |
| Corticosteroids | 20 (17.2) | 52 (16.6) |  |
| Colchicine | 8 (6.8) | 14 (4.5) |  |
| CVVDHF | 1 (0.9) | 4 (1.3) |  |
| **Final Outcome** |  |  |  |
| Survival | 93 (79.5) | 233 (74.0) | 0.256 |
| Death | 24 (20.5) | 81 (26.0) |  |
| **Sampling approach** |  |  |  |
| Single | 40 (34.2) | 40 (12.7) | <0.001 |
| Multiple | 77 (65.8) | 274 (87.3) |  |
| **Day from disease onset at sampling** |  |  |  |
| Day 1-7 | 37 (31.6) | 55 (17.5) | 0.004 |
| Day 8-14 | 43 (36.8) | 124 (39.5) |  |
| Day 15+ | 37 (31.6) | 135 (43.0) |  |
| **Follow-up period** |  |  |  |
| Median ($\pm 95\%$ CI) | 21.0±2.1 | 22.0±2.3 | 0.070 |
| **Hospital** |  |  |  |
| Alexandroupolis | 68 (58.1) | 194 (61.8) | 0.488 |
| AHEPA | 49 (41.9) | 120 (38.2) |  |

*CVVDHF: continuous venovenous hemodiafiltration; Corticosteroids: dexamethasone or equivalent doses of alternative glucocorticoids; LMWH: low molecular weight heparin*

### Immunoassays

Activin-A, Activin-B and Follistatin serum levels were measured using enzyme-linked immunosorbent-assay (ELISA) (Ansh-Labs, Webster, TX, USA) according to manufacturer’s instructions.

### Statistical analysis

To elucidate the role of A/F-axis molecules, we identified potent confounders from true predictors of outcome using general linear model. Prognostic value of predictors was validated; for that purpose, all independent variables of interest were transformed to binary ones through nominal optimal scaling along with discretization to two groups. Imputing of missing data was added, ridge-regression was selected for regularization and 10-fold cross-validation was added. Results were adapted to a point-system scoring, where one additive point was given for each parameter included in the unfavorable category as suggested after discretization; binary-regression was utilized to mathematically approach outcome probability. Evaluation of the scoring systems was based on Area-Under-Curve (AUC) as determined from Receiver Operating Characteristic (ROC) analysis. Optimal-Scaling procedure was utilized to detect whether the scoring systems could predict response to certain treatment options. More details regarding the Materials and Methods is provided in Supplementary Materials.

## Results

### The Activin/Follistatin axis is highly deregulated in COVID-19 non-survivors

To validate the hypothesis that the A/F-axis is linked to COVID-19 pathophysiology we analyzed serum levels of Activin-A, Activin-B and Follistatin in a cohort of 117 COVID-19 patients. The day-of-symptom-onset (disease-day) was used to align the data derived from serum samples. Activin-A, Activin-B and Follistatin, were substantially increased, particularly in samples from non-survivors (Figures 1 and 2, and Supplementary Table 2). Increased levels of A/F-axis components were observed approximately 7-28 days from onset of symptoms (Figures 1, 2). Classical parameters such as ANC, ALC, NLR, CRP, LDH and D-dimers became derailed within the same time window (Figure 1). However, whereas CRP, neutrophils and NLR were elevated already from first week of disease and D-dimers were gradually increasing over time, A/F-axis components and LDH were upregulated during the second week, when COVID-19 leads to hospitalization (Figure 2). Interestingly, comparative analysis of A/F-axis components and classical parameters in survivors *vs* non-survivors on the basis of disease-status at serum sampling (Supplementary Figure 2) demonstrated upregulation of Activin-A, Activin-B and Follistatin in sera from both severe and critical, disease status (DS) 3 and DS4, non-survivors. Neutrophils, CRP and D-dimers were upregulated in samples from DS3 non-survivors, as well as, DS4 survivors and non-survivors; LDH levels were particularly elevated in DS4 non-survivors.

**Figure 1.**
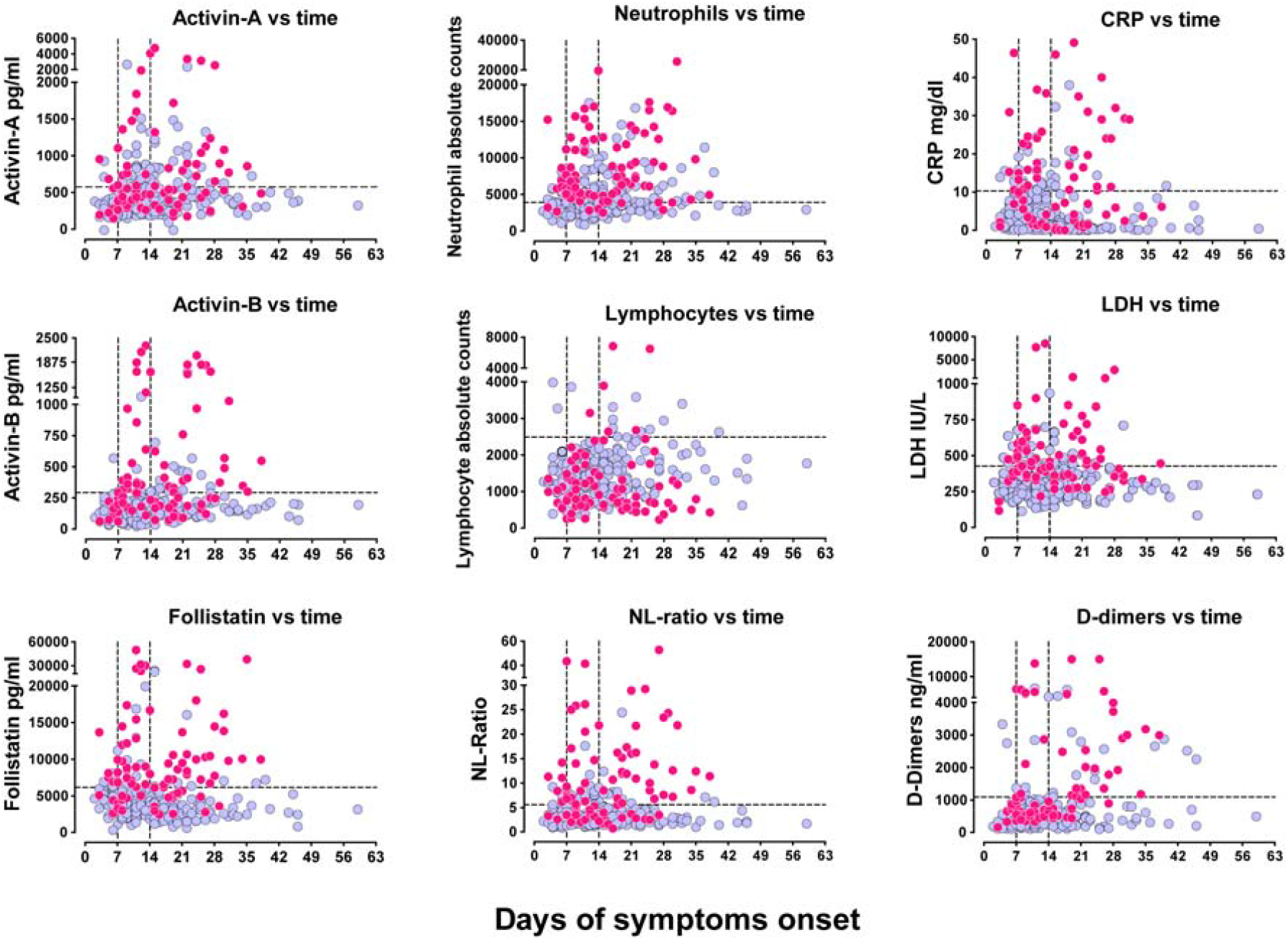
Activin/Follistatin-axis components are upregulated in the serum of COVID-19 patients during the critical period when patients deteriorate. Serum levels of A/F-axis components and other key inflammatory parameters in the blood of COVID-19 patients plotted over time according to the days of symptom onset. Light blue circles represent samples collected from patients that survived COVID-19 and magenta circles represent samples obtained from patients that eventually succumbed to the disease. Vertical dotted lines indicate time-windows of first, second and more than two-week intervals used for statistical analysis. Horizontal dotted lines indicate threshold as determined by the nominal optimal scaling along with discretization to two groups.

**Figure 2.**
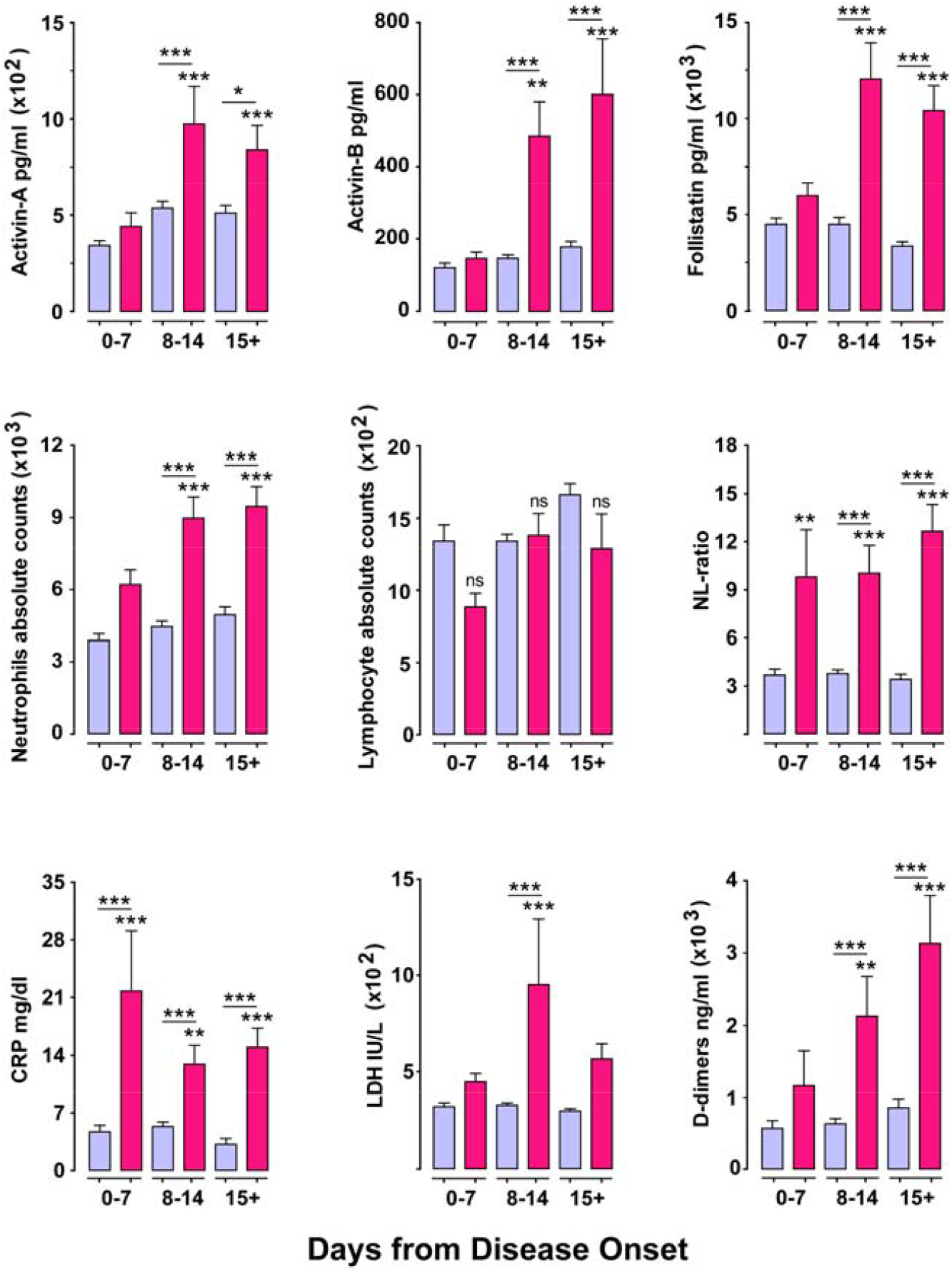
Levels of A/F-axis components are upregulated selectively in non-survivor severe or critically-ill COVID-19 patients during the critical second week from disease onset. Concentrations of Activin/Follistatin-axis components in serum and levels of other key inflammatory parameters in the blood of COVID-19 patients plotted over time according to the days of symptom onset and grouped per week. Light blue bars represent samples collected from patients that survived COVID-19 and magenta bars represent samples obtained from patients that eventually succumbed to the disease. Data are expressed as mean±SEM analyzed using one-way analysis of variance with Tuckey’s post-hoc test. Asterisks represent comparison of the indicated group vs the survivors of week 1 (0-7 days from onset of disease). Asterisks above a horizontal line represent comparison between the groups under the line segment (*P<0.05, **P< 0.01, and ***P < 0.001)

To assess whether sampling (samples/patient, disease-day at sampling) and origin of patients (hospital of admission) might blur the results, a general linear model including Activin-A, Activin-B and Follistatin as dependent variables and multiplicity of samples/patient, sampling period (1-7/8-14/15+ days) and reference hospital as independent were introduced. The model demonstrated that Activins and Follistatin were not correlated with these potent confounders (Supplementary Table 3). Therefore, our efforts to construct a predictive model were sample-based rather than patient-based.

### The Activin/Follistatin-axis in the dynamically shifting phenotypic heterogeneity of COVID-19

COVID-19 is characterized by clinical heterogeneity and a dynamically changing phenotype [2,3,5,12]. We hypothesized that this behavior must be mirrored by analogous fluctuations in the expression profiles of key biomarkers. We therefore investigated the correlations between different biomarkers and final outcome in our sample collection. The correlation of Activin-B levels to LDH, and Activin-A levels to CRP and NLR are shown as representative comparisons (Figure 3). In Activin-B vs LDH, some samples exhibited good correlation but others did not and in Activin-A vs CRP or NLR comparisons, the parameters did not correlate at all. To understand better this finding, we highlighted in similar plots the groups of samples derived from non-survivors AL022, AL034, AL035, AL062, AL063 and AX112 for which multiple samplings were available (Figure 3). Remarkably, this depiction highlighted the marked and dynamic phenotypic heterogeneity among patients and samples of the same patient collected at different disease-days. For example, for patients AL022 and AL035, disease progression and eventual death were associated with concordant changes in Activin-B and LDH levels. However, patients AL063 and AX112, were characterized by substantial changes in CRP and A/F-axis component levels and for patients AL035 and AL062 disease progression was associated with eventual CRP and NLR reduction. Interestingly, patient AL022, who received a dose of Tocilizumab one day after admission, exhibited for ∼10 days transient reduction of LDH, CRP, NLR, Activin-A and Activin-B, before reverting and dying on disease-day 28. Therefore, in addition to patient intrinsic phenotypic variability, treatments could also influence biomarker profile during hospitalization. From all the above we concluded that a broader spectrum of biomarkers, including both early and later modulated ones, had to be integrated to develop a meaningful disease scoring/monitoring-system.

**Figure 3.**
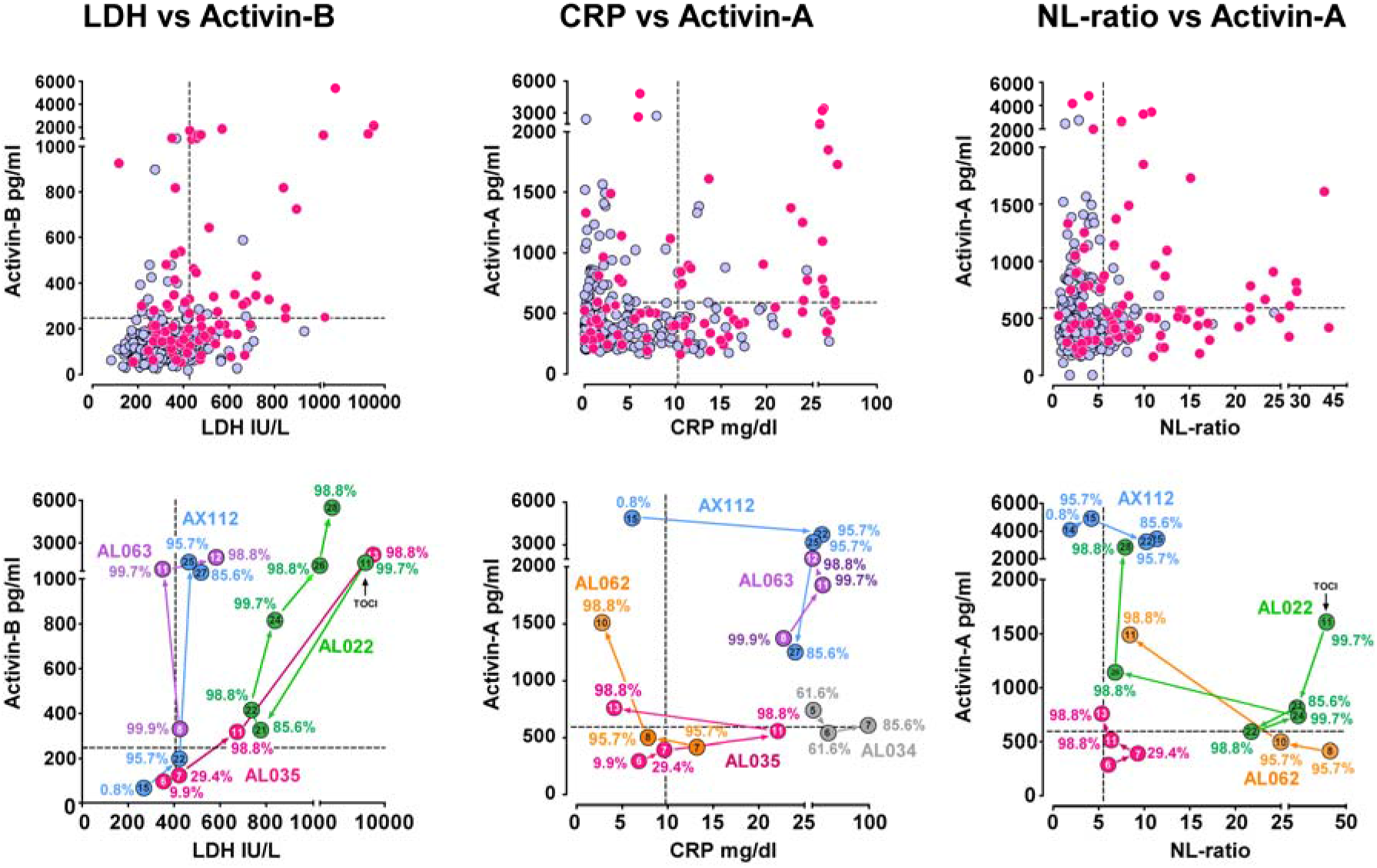
The dynamic phenotypic heterogeneity of COVID-19 patients is evident when serum relative levels of A/F-axis components are plotted against classical biomarkers. Upper panel: Correlation between Activin-B and LDH or Activin-A and CRP or NL-ratio levels in the randomly acquired serum samples from our cohort. Vertical and horizontal dotted lines indicate the thresholds as determined by the nominal optimal scaling along with discretization to two groups. Lower panel: Similar correlations as above, however, serum samples derived from the same patient are indicated with distinct colors and connected with arrows to indicate the direction of disease progression towards death. Numbers within the circular points indicate disease-day. Percentages indicate the probabilities of death estimated by the FACT-CLINYCoD scoring system developed herein. TOCI: Tocilizumab

### The FACT-CLINYCoD score for dynamic monitoring of COVID-19 outcome and treatment

Considering the dynamic and heterogeneous phenotypic changes characterizing COVID-19 progression, we aimed to build a simple and clinically meaningful scoring system to discriminate survivors from non-survivors. To this end, we first demonstrated that Follistatin, Activin-A, Activin-B, CRP, LDH, ICU-admittance, NLR, Age, comorbidity numbers and D-dimers performed well in predicting disease outcome, as judged by ROC analysis (Supplementary Figure 3). Notably, among all these parameters, Follistatin exhibited the highest AUC (0.857). We then converted all these continuous variables, which could independently predict outcome, to binary through selection of optimal cutoffs using optimal-Scaling procedure along with ridge-regression (Supplementary Figure 4 and Supplementary Table 4). Based on these data, we constructed a ten-point scoring-system where one additive point was arbitrarily given for each parameter included in the unfavorable category as suggested after discretization, namely if Follistatin>6235 pg/ml, Activin-A>591 pg/ml, Activin-B>249 pg/ml, CRP>10.3 mg/dL, LDH>427 U/L, ICU admission, NLR>5. 6, Years of Age >61, Comorbidities>1 and D-dimers>1097 ng/ml (Table 2, Supplementary Table 4). We evaluated this score with ROC analysis, yielding to an AUC value of 0.951±0.032 (p<10^−6^), that indicates outstanding discrimination in foretelling survivors from non-survivors (Figure 4A). This score was titled FACT-CLINYCoD, being an acronym of *F*(ollistatin), *ACT*(ivins), *C*(RP), *L*(DH), *I*(CU admission), *N*(LR), *Y*(ears of age), *Co*(morbidities) and *D*(-dimers). A score ≥4 has 90.8% sensitivity and 87.5% specificity to predict fatal outcome, whereas a score ≥5 has 81.5% sensitivity and 95.7% specificity. The FACT-CLINYCoD score distinguishes survivors from non-survivors at admission and outcome and monitors accurately disease progression independently of disease-day or status (Figure 4B,C,D). The probabilities of death at different disease-days in non-surviving patients AL022, AL034, AL035, AL062, AL063 and AX112 are shown in Figure 3, next to each sample analyzed. Interestingly, some of these patients had very high scores already at first sampling, however, others, such as AX112 and AL035 had low scores at admission and either gradually or rapidly deteriorated and eventually died. The latter type of patients highlights the value of continuous monitoring for COVID-19.

**Table 2.**
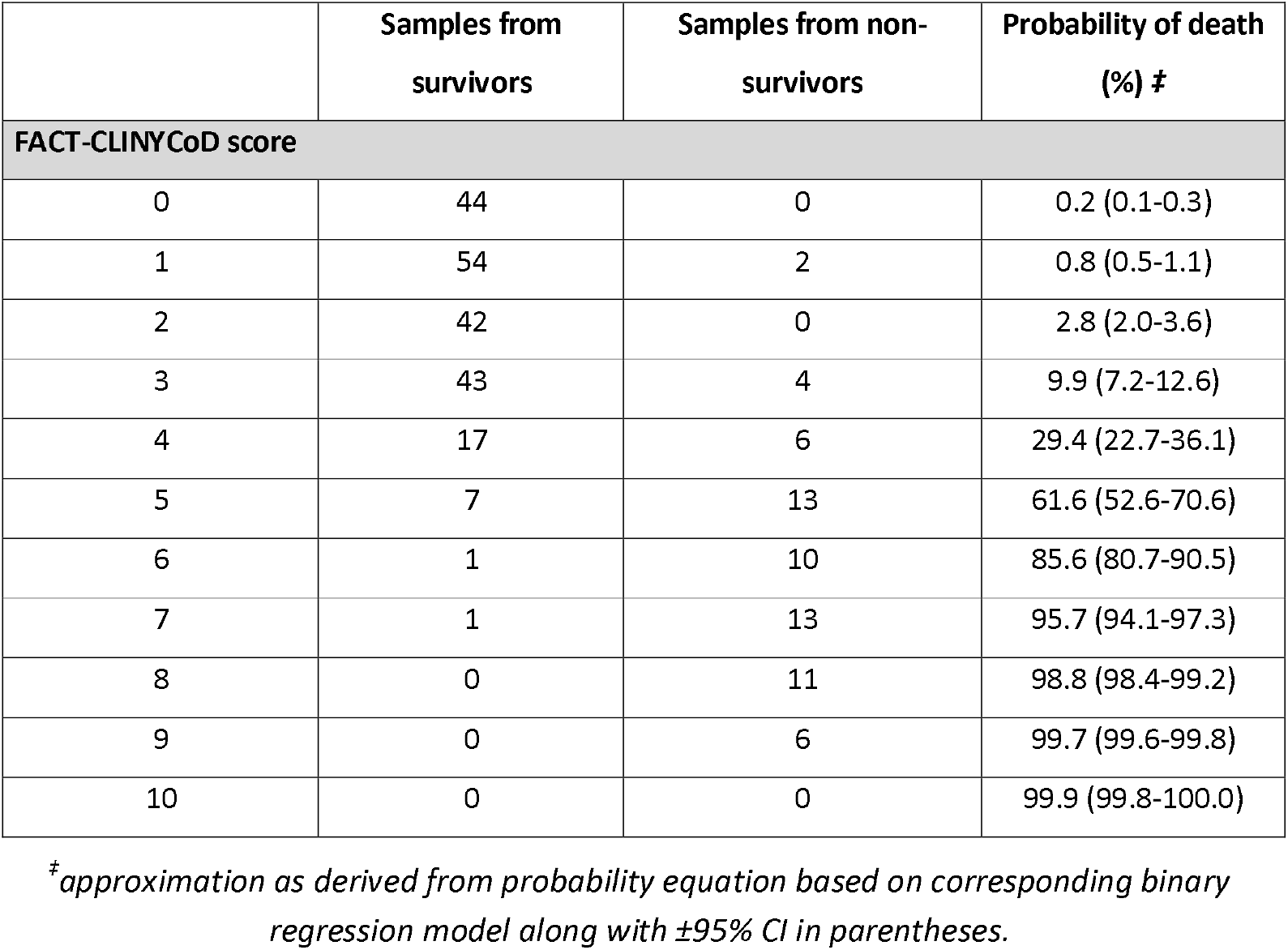
FACT-CLINYCoD scoring and predictability of outcome.

|  | Samples from survivors | Samples from non-survivors | Probability of death (%) <sup>‡</sup> |
| --- | --- | --- | --- |
| **FACT-CLINyCoD score** |  |  |  |
| 0 | 44 | 0 | 0.2 (0.1-0.3) |
| 1 | 54 | 2 | 0.8 (0.5-1.1) |
| 2 | 42 | 0 | 2.8 (2.0-3.6) |
| 3 | 43 | 4 | 9.9 (7.2-12.6) |
| 4 | 17 | 6 | 29.4 (22.7-36.1) |
| 5 | 7 | 13 | 61.6 (52.6-70.6) |
| 6 | 1 | 10 | 85.6 (80.7-90.5) |
| 7 | 1 | 13 | 95.7 (94.1-97.3) |
| 8 | 0 | 11 | 98.8 (98.4-99.2) |
| 9 | 0 | 6 | 99.7 (99.6-99.8) |
| 10 | 0 | 0 | 99.9 (99.8-100.0) |

<sup>‡</sup>*approximation as derived from probability equation based on corresponding binary regression model along with $\pm 95\%$ CI in parentheses.*

**Figure 4.**
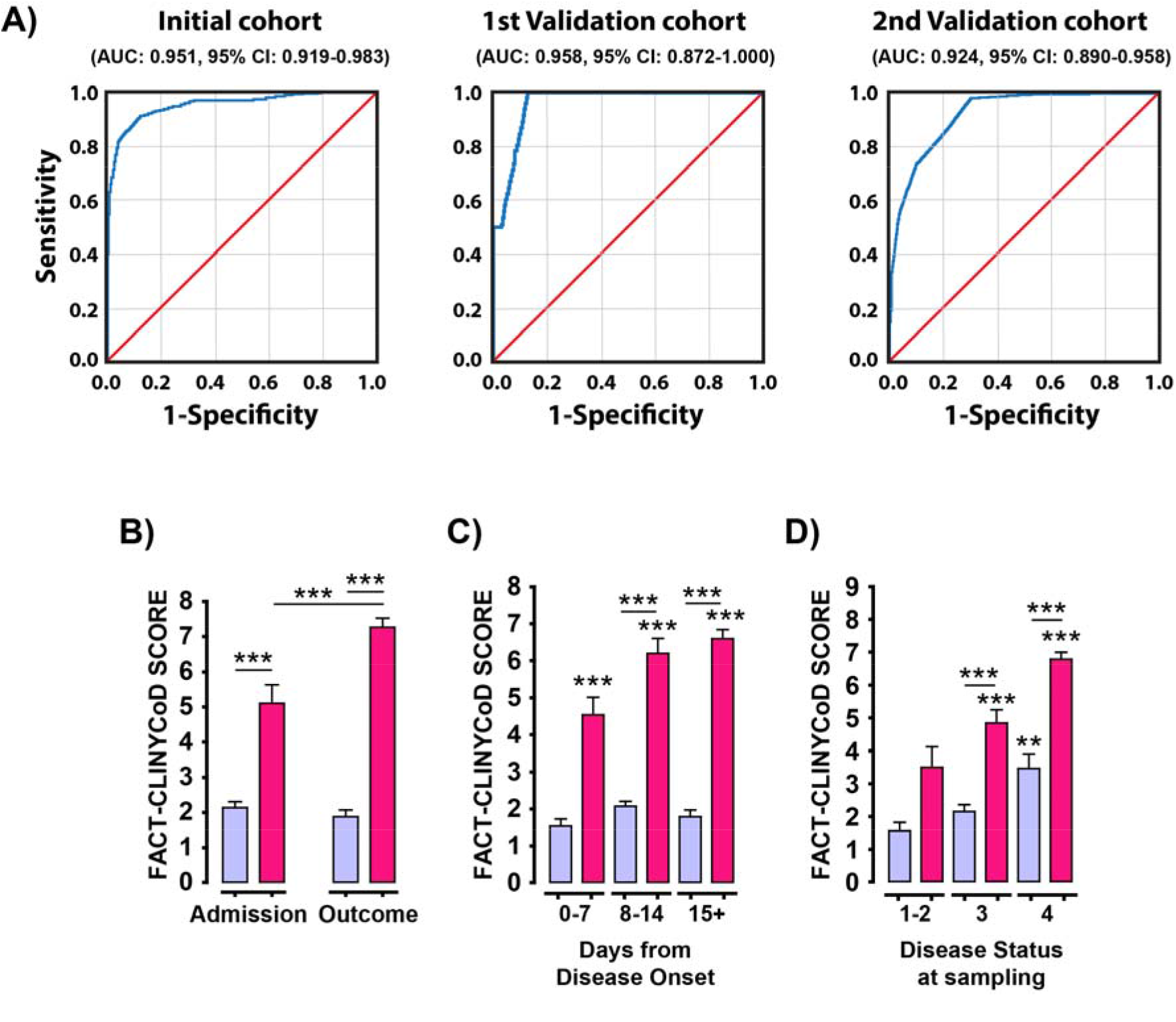
The FACT-CLINYCoD scoring system may enable prediction of day-to-day prognosis of disease outcome and real-time clinical decision making. ROC analysis for FACT-CLINYCoD score of the initial and two validation cohorts (A). The FACT/CLINYCoD scoring system distinguishes survivors from non-survivors both at admission and at the final outcome (B), irrespectively of disease-day (C) and disease status (D). Light blue bars represent samples collected from patients that survived COVID19 and magenta bars represent samples obtained from patients that eventually succumbed to the disease. Data are expressed as mean±SEM analyzed using one-way analysis of variance with Tuckey’s post-hoc test. Asterisks above a horizontal line represent comparison between the groups under the line segment. Solitary asterisks in C and D represent comparison of the indicated group with the survivors in the first group of the corresponding graph (*P<0.05, **P< 0.01, and ***P < 0.001).

Considering that various treatment modalities might affect the value of FACT-CLINYCoD score to predict final outcome, we performed ROC analysis separately for samples corresponding to each treatment (Supplementary Figure 5). Interestingly, all relevant AUCs were comparable, thus indicating that the predictive value of FACT-CLINYCoD score was not affected by current therapies. Therefore, this scoring system may be used for monitoring response to treatment.

### Validation of the FACT-CLINYCoD score during the second wave of COVID-19

The FACT-CLINYCoD score was at first validated using an independent cohort of 28 consecutive patients contributing 35 random samples. Although this small cohort differed significantly from the initial regarding mortality rate, age, disease-status, treatments, sampling approach, and day-from-disease-onset (Supplementary Table 1), a comparable AUC was observed (0.958, 95%CI: 0.872-1.000, P=0.032, Figure 4A). A second independent validation cohort of 118 consecutive patients contributing 225 samples, differing significantly regarding sex-ratio, number of comorbidities, disease-status, treatments, and sampling approach (Supplementary Table 1) was again found to exhibit an AUC of 0.924 (95%CI: 0.890 - 0.958, P<10^−6^, Figure 4A). Both validation cohorts shared comparable AUCs with the initial cohort (P=0.880 and 0.256, respectively).

### The value of FACT-CLINYCoD score to guide timely treatment

Optimal-Scaling procedure was performed along with ridge-regression to investigate whether FACT-CLINYCoD score could guide timely administration of current SOC treatments [25] such as low-molecular-weight heparin (LMWH), remdesivir and dexamethasone (or equivalent doses of alternative glucocorticoids) on data available from 574 samples after pooling the initial and validation cohorts. Both LMWH (P<0.001) and dexamethasone (P=0.026), when administered in FACT-CLINYCoD score ≤2 (P<0.001), were independently correlated with favorable outcome. This was not the case for remdesivir (P=0.071), most possibly due to the limited number of available samples (Supplementary Table 6). Although the utility of FACT-CLINYCoD in guiding timely treatment administration and predicting therapeutic responses looks promising, the relatively small number of patients dictates caution in interpreting these findings.

## Discussion

This study provides evidence suggesting that inflammation and tissue stress-related proteins Activin-A, Activin-B, and Follistatin (A/F-axis) are tightly associated with severity and outcome of COVID-19. Their upregulation was prominent in non-survivors and was independently related to in-hospital mortality. Of note, combination of A/F-axis components with common clinical and laboratory parameters permits prediction of COVID-19 outcome throughout the course of the disease.

Identification of high-risk patients and death-prediction are of particular importance especially in strained health-care systems. Hence, several studies attempted to develop scoring systems to predict timely disease severity and mortality [4,6,14,15,26–28]. Most of these models were based on demographic parameters and instantaneous evaluation of subjective symptoms (i.e. dyspnea) combined with single measurements of common laboratory markers, such as neutrophil or lymphocyte counts, LDH, CRP and several proinflammatory cytokines [4,6,14,15,26,28,29]. These models are undoubtedly useful; however, they emphasize prediction of final-outcome at admission and thus may fail to predict some non-survivors most likely due to the complex and often erratic development of COVID-19 and the downregulation of key biomarkers at late disease-stages (Figure 3).

Considering the above, we exploited the findings presented herein to develop a more dynamic COVID-19 monitoring-system. Specifically, we utilized the measurement of Activin and Follistatin levels, based on their deregulation described herein and their previous implication in sepsis/ARDS, neutrophil-mediated inflammation, coagulopathy, endothelial cell stress, angiogenesis and post-inflammatory pulmonary fibrosis/remodeling, all of which are key characteristics of COVID-19 pathophysiology [7,8,18,22,30]. Moreover, we reasoned that upregulation of A/F-axis could indicate tissue-damage in the lung, vasculature or other vital peripheral tissues, thus complementing information derived from other biomarkers reporting tissue-damage (i.e. LDH) or coagulation/vascular injury (i.e. D-dimers) [2,15,27,28]. Finally, taking into consideration the differences in kinetics of A/F-axis components and other clinical biomarkers like NLR, CRP, LDH and D-dimers (Figure 2) we combined them all to develop a more generalizable monitoring-system that will not only make early predictions but will rather allow readjustment of predictions during disease progress [31].

In the pre-COVID-19 era, the A/F-axis was evaluated as predictor of outcome in critical-care patients [32]. Increased levels of Activin-A and Activin-B in sera from ICU patients with acute respiratory failure could predict 90-days and 12-months survival with reasonable accuracy (∼80%). Follistatin did not provide any extra predictive value [32]. A/F-axis components were measured also in critically-ill, influenza-A (H1N1), patients, but no significant association with disease severity was established [33]. However, while preparing the current manuscript, consistent with our conclusions, a preprint study reported Follistatin among the circulating markers of endothelial damage associated with in-hospital mortality in a small number of COVID-19 patients [34].

Although the exact mechanism linking Activins to COVID-19 pathogenesis remains unknown, the importance of A/F-axis in the disease is reflected on the predictive value of FACT-CLINYCoD score. This model, which depends on A/F-axis by 3/10, demonstrates an almost perfect AUC (0.951±0.032) as well as a very satisfactory sensitivity (81.5%) and specificity (95.7%) of scores ≥5 to predict fatal outcome. In agreement with this, the key role of A/F-axis components, in particular Follistatin, in the prediction of fatal outcome at any time of disease course (Figure 4B,C) is underlined by the relevant ROC analysis (Supplementary Figure 3) and ridge-regression (Supplementary Figure 4). The FACT-CLINYCoD score constitutes a balanced, robust and flexible tool efficiently intertwining pathophysiology of A/F-axis, clinical profile (by means of NLR, LDH, D-dimers and CRP) and key parameters that are well established to affect mortality (ICU admission, age, and comorbidities), capable of providing dynamic outcome prediction.

An obvious emerging question is whether the A/F-axis is a suitable target for COVID-19 therapeutics. Indeed, several studies have proposed Follistatin or soluble Activin type-II receptors as therapeutics for sepsis, ARDS and fibrotic disorders [18,35]. Although increase of Activin-A and -B could be interpreted in favor of utilizing such therapeutics, the dramatic increase of Follistatin, often at stoichiometry that surpasses substantially the sum of Activins-A and -B in serum, could argue against it. Neutrophils can release preformed Activin-A upon arrival in inflamed tissues [18,36], whereas, other inflammatory cells such as monocytes, CD4^+^ T cells and tissue resident cells can secrete Activin-A later on [18,21,37]. Neutrophils do not secrete Activin-B and therefore this factor must be produced by other cells, probably acting as a biomarker of vascular stress and hypoxia [38,39]. Follistatin can be produced locally or released systemically from distant organs such as liver [18]. The tissue origin of the increased A/F-axis components in sera of COVID-19 patients and the actual Activin/Follistatin stoichiometry in the affected tissues are unknown. Therefore, the suitability of the A/F-axis as therapeutic target for COVID-19 warrants further careful investigation.

Another promising aspect of this study is the potential utility of FACT-CLINYCoD to monitor response to various treatments targeting SARS-CoV-2 (antivirals) or hyper-inflammatory host reactions (heparin, corticosteroids) since the value of this scoring system to predict mortality was not disturbed by various treatments. Our preliminary results showed favorable outcome when LMWH and dexamethasone was commenced at FACT-CLINYCoD score ≤2. This is consistent with emerging clinical data linking reduced mortality with early administration of LMWH in all hospitalized COVID-19 patients [40] and supports the detrimental role of immunothrombosis in COVID-19 [7]. Moreover, current clinical data derived from randomized clinical studies recommend administration of dexamethasone as early as the patient needs oxygen supply [25].

Finally, the postulated implication of the A/F-axis in other infectious diseases associated with in-hospital mortality [17–19,22], leaves open the possibility that the FACT-CLINYCoD score may be applicable to other diseases in addition to COVID-19. In conclusion, A/F-axis dysregulation is tightly associated with poor outcome of COVID-19. FACT-CLINYCoD, a novel pathophysiology-driven monitoring-system, enables dynamic prediction of disease outcome and may support real-time medical decision. Prospectively, large-scale, multinational validation of this calculator, as well as investigation of the mechanisms linking A/F-axis to COVID-19 pathogenesis is definitely warranted.

## Data Availability

All data referred to in the manuscript are avaliable upon request.

## Acknowledgements

We thank Dr. Apostolos Vasileiou for technical assistance and Dr. Mika Laitinen for critical reading of the manuscript.

## Supplementary Data

### Study criteria and disease status

Inclusion criteria of the study were: a) adult patients (>18 years old), any gender; b) positive SARS-CoV-2 RT-PCR testing in nasopharyngeal swab or BAL; c) hospitalization due to COVID-19, any disease stage; d) Known final disease outcome. Patients without available sample obtained before the 21^st^ day-of-disease were excluded.

Comorbidities considered were diabetes mellitus, arterial hypertension, dyslipidemia, obesity, atrial fibrillation, coronary disease, heart-failure, renal-failure, chronic obstructive pulmonary disease or asthma, immunosuppression (without a record of malignancy), autoimmunity and cancer.

The disease-status (DS) of COVID-19 patients was classified based on the adaptation of the Sixth Revised Trial Version of the Novel Coronavirus Pneumonia Diagnosis and Treatment Guidance, as described previously by Hadjadj et al [1]. Specifically, mild cases (DS1) were defined as mild clinical symptoms (fever, myalgia, fatigue, diarrhea) and no sign of pneumonia on thoracic X-Ray or/and CT scan. Moderate cases (DS2) were defined as clinical symptoms associated with dyspnea and radiological findings of pneumonia on thoracic X-Ray or/and CT scan, and requiring a maximum of 3 L/min of oxygen. Severe cases (DS3) were defined as respiratory distress requiring more than 3 L/min of oxygen and no other organ failure. Critical cases (DS4) were defined as respiratory failure requiring mechanical ventilation, shock and/or other organ failure that require an intensive care unit (ICU).

### Immunoassays

Activin-A, -B and Follistatin levels in serum were measured using enzyme-linked immunosorbent assay (ELISA) (Ansh Labs, Webster, TX, USA) according to manufacturer’s instructions. For Activin-A (Cat# AL-110) limit of detection was 65 pg/ml and dynamic range 0.1-10 ng/ml. For Activin-B (Cat# AL-150) limit of detection was 4.35 pg/ml and the dynamic range 12.7-1400 pg/ml. For Follistatin (Cat# AL-117) limit of detection was 180 pg/ml and dynamic range 0.612-20 ng/ml. Baseline levels of Activin-A, -B and Follistatin in 22 normal, pre-COVID-19 sera, were 405±180, 145±54 and 2607±1665 pg/ml, respectively, very similar to those previously reported by others using the same reagents [2]. The investigators who performed the immunoassays were blinded towards any data that could serve to identify patients, such as names, addresses, and social insurance ID, in accordance to the TRIPOD statement [4].

### Statistics

To search for biases, we compared all measured parameters between patients and samples using either Pearson’s χ^2^ or a two-sample independent t-test in case of discrete and continuous variables respectively; in case of statistically significant increase in homogeneity of variance, as checked by Levene’s test, Mann-Whitney U test was additionally performed. General linear model was used to dissect true independent correlations of the A/F axis molecules from potent confounders. All initially selected variables (LDH, CRP, NLR, D-dimers, ANC, ALC, Ferritin, Age, Comorbidities, ICU admission, Follistatin, Activin-A, and Activin-B, multiplicity of sampling, period of sampling, and reference hospital) were transformed to binary ones through nominal optimal scaling along with discretization to two groups, imputing of missing data was added, and 10-fold cross-validation was selected through SPSS CATREG procedure. Models including parameters with tolerance <0.4 (variance inflation factor >2.5), were rejected to avoid collinearity. Furthermore, ridge regression was used to avoid model overfitting, tolerate large variances and overcome collinearity obstacles, all at the least possible additive bias. However, aiming to ascertain further interpretation and adaptation to reach clinical usefulness, we constructed and evaluated a point-system scoring incorporating all parameters with independent prediction validity (Follistatin, Activin-A, and Activin-B, CRP, LDH, ICU admission, NLR, Age, Comorbidities, D-dimers). One additive point was arbitrarily given for each parameter included in the unfavorable category as suggested after discretization; to assess correlation of final score with outcome, binary regression was utilized. The accuracy of the scoring system was approached using the Area Under Curve (AUC) as determined from Receiver Operating Characteristic (ROC) analysis. Optimal Scaling procedure was utilized as described before to detect whether the scoring systems could predict response to certain treatment options.

Using binary regression analysis by assessing numerical values to outcome, the obtained probability of unfavorable outcome (death) was given by the formula p=1/(1+*e*^*-Π*^), where Π stands for constant + ∑b_n_x_n_; 95% CI of p lie between exp[Π-∑1.96* SE(b_n_)]/{1+exp[Π-∑1.96*SE(b_n_)]} and exp[Π-∑1.96*SE(b_n_)]/{1+exp[Π+∑1.96* SE(b_n_)]}.

Estimation of the minimum sample size for both survivors and non-survivors corresponding to an AUC>0.750 with a<0.05 and b<0.20 as well as evaluation of the difference between independent ROC curves was performed with the use of MedCalc Statistical Software version 19.4.1 (MedCalc Software Ltd, Ostend, Belgium; https://www.medcalc.org; 2020). Imputed data tolerated missing values at a maximum of 5%, to confirm transparency and generalizability [3]. The level of statistical significance was set to p=0.05. All numerical values are given with at least two significant digits. Means are accompanied by their 95% confidence intervals (CI). Statistical analysis was performed with the use of IBM SPSS Statistics software, version 26.0.0.0, for Windows. The investigators who analyzed data and built the prediction model were blinded towards any data that could serve to identify patients, such as names, addresses, and social insurance ID, to ensure that the prediction model would fully conformed to the TRIPOD statement [4].

### The FACT-CLINYCoD score for COVID-19 outcome prognosis

Optimal Scaling procedure along with ridge regression yielded to a 10-parameter model as presented in Table 2, Figure 4, and Supplementary Figure 4. Based on these data, we constructed a ten point-system scoring where one additive point was arbitrarily given for each parameter included in the unfavorable category as suggested after discretization, namely if Follistatin >6235 pg/ml, Activin-A >591 pg/ml, Activin-B >249 pg/ml, CRP >10.3 mg/dL, LDH >427 U/L, ICU admission, NLR >5.6, Age >61 years, Comorbidities >1 and D-dimers >1097 ng/ml.

A binary regression model incorporating outcome as a dependent variable and score as an independent one (Supplementary Table 5) yields to the formula p=1/(1+*e*^6.198 – 1.330*score^), where p stands for the probability of death at the time of sampling; ±95% CI of p lie between e^-6.547+1.330*score^ /(1+e^-6.547+1.330*score^) and e^-5.849+1.330*score^ /1+e^-^).^5.849+1.330*score^

## SUPPLEMENTARY TABLES

**Supplementary Table 1.** Characteristics of samples derived from first (n=35) and second (n=225) validation cohort in comparison with samples derived from initial analysis cohort (n=314).

| Parameter | Initial cohort (n=314) | First validation cohort (n=35) | Second validation cohort (n=225) |
| --- | --- | --- | --- |
| **Age (years)** |  |  |  |
| Mean ($\pm 95\%$ CI) | 61.6±1.7 | 71.2±5.9‡ | 59.9±2.1 |
| **Sex** |  |  |  |
| Male | 161 (51.3) | 19 (54.3) | 138 (61.3)† |
| Female | 153 (48.7) | 16 (45.7) | 87 (38.7)† |
| **Number of comorbidities** |  |  |  |
| 0 | 69 (22.0) | 3 (8.6) | 73 (32.4)† |
| 1 | 79 (25.2) | 15 (24.9) | 65 (28.9)† |
| 2 | 72 (22.9) | 7 (20.0) | 37 (16.4)† |
| 3 | 60 (19.1) | 6 (17.1) | 34 (15.1)† |
| 4 | 28 (8.9) | 4 (11.4) | 9 (4.0)† |
| >5 | 6 (1.9) | 0 (0.0) | 7 (3.1)† |
| **Disease status (DS) at sampling** |  |  |  |
| DS1 | 95 (30.2) | 24 (68.6)‡ | 47 (20.9)‡ |
| DS2 | 74 (23.6) | 5 (14.3)‡ | 83 (36.9)‡ |
| DS3 | 71 (22.6) | 1 (2.9)‡ | 50 (22.2)‡ |
| DS4 | 74 (23.6) | 5 (14.3)‡ | 45 (20.0)‡ |
| **Treatment at sampling** |  |  |  |
| Hydroxychloroquine | 98 (31.2) | 26 (74.3)‡ | 56 (24.9) |
| Azithromycin | 125 (39.8) | 21 (60.0)† | 85 (37.8) |
| Antibiotics (other) | 257 (81.8) | 12 (34.3)‡ | 173 (76.9) |
| Lopinavir/Ritonavir | 112 (35.7) | 0 (0.0)‡ | 34 (15.1)‡ |
| Remdesivir | 10 (3.2) | 0 (0.0) | 4 (1.8) |
| Tocilizumab | 5 (1.6) | 0 (0.0) | 10 (4.4)† |
| Anakinra | 31 (9.9) | 0 (0.0) | 54 (24.0)‡ |
| LMWH | 254 (80.9) | 15 (42.9)‡ | 213 (94.7)‡ |
| Corticosteroids | 52 (16.6) | 0 (0.0)‡ | 116 (69.0)‡ |
| Colchicine | 14 (4.5) | 0 (0.0) | 9 (4.0) |
| CVVDHF | 4 (1.3) | 0 (0.0) | (0.0) |
| **Sampling approach** |  |  |  |
| Single | 40 (12.7) | 21 (60.0)‡ | 48 (21.3)‡ |
| Multiple | 274 (87.3) | 14 (40.0)‡ | 177 (78.7)‡ |
| **Day from disease onset at sampling** |  |  |  |
| Day 1-7 | 55 (17.5) | 14 (40.0)‡ | 51 (22.6) |
| Day 8-14 | 124 (39.5) | 13 (37.1)‡ | 87 (38.7) |
| Day 15+ | 135 (43.0) | 8 (22.9)‡ | 87 (38.7) |
| **Follow-up period** |  |  |  |
| Median ($\pm 95\%$ CI) | 22.0±1.3 | 30.0±8.6 | 22.0±1.4 |
| **Final Outcome** |  |  |  |
| Survival | 233 (74.0) | 32 (91.4)† | 168 (74.7) |
| Death | 81 (26.0) | 3 (8.6)† | 57 (25.3) |
| **Hospital** |  |  |  |
| Alexandroupolis | 194 (61.8) | 0 (0.0)‡ | 102 (45.3)‡ |
| AHEPA | 120 (38.2) | 0 (0.0)‡ | 123 (54.7)‡ |
| Attikon | 0 (0.0) | 35 (100.0)‡ | 0 (0.0) |

CVVDHF: continuous venovenous hemodiafiltration; Corticosteroids: dexamethasone or equivalent doses of alternative glucocorticoids; LMWH: low molecular weight heparin; $\dagger 0.05 < P \leq 0.01$ vs initial cohort; $\ddagger P < 0.01$ vs initial cohort

**Supplementary Table 2.** Comparison between survivors and non-survivors in the initial cohort (samples).

| Parameter | survivors (n=231) | non-survivors (n=81) | P |
| --- | --- | --- | --- |
| **Sample number per patient** |  |  |  |
| Mean ($\pm 95\%$ CI) | 3.5±0.2 | 5.2±0.6 | <0.001 |
| **Collection day per sample** |  |  |  |
| Mean ($\pm 95\%$ CI) | 14.2±1.1 | 16.4±1.8 | 0.043 |
| **Hospital** |  |  |  |
| Alexandroupolis | 132 (57.1) | 60 (74.1) | 0.007 |
| AHEPA | 99 (42.9) | 21 (25.9) |  |
| **Age (years)** |  |  |  |
| Mean ($\pm 95\%$ CI) | 57.4±2.0 | 73.2±2.0 | <0.001 |
| **Sex** |  |  |  |
| Male | 112 (48.5) | 39 (48.1) | 0.958 |
| Female | 119 (51.5) | 42 (51.9) |  |
| **Comorbidities (n)** |  |  |  |
| 0 | 62 (26.8) | 7 (8.8) | <0.001 |
| 1 | 68 (29.4) | 11 (13.8) |  |
| 2 | 61 (26.4) | 11 (13.8) |  |
| 3 | 27 (11.7) | 31 (38.8) |  |
| 4 | 9 (3.9) | 19 (23.8) |  |
| 5 | 5 (2.2) | 1 (1.3) |  |
| **Disease status (DS) at sampling** |  |  |  |
| DS1 | 94 (40.7) | 1 (1.3) | <0.001 |
| DS2 | 69 (29.9) | 5 (6.3) |  |
| DS3 | 52 (22.5) | 17 (21.3) |  |
| DS4 | 17 (7.4) | 57 (71.3) |  |
| **Treatment** |  |  |  |
| Hydroxychloroquine | 77 (33.5) | 21 (25.9) | 0.061 |
| Azithromycin | 92 (40.0) | 31 (38.3) |  |
| Antibiotics (other) | 184 (80.0) | 71 (87.7) |  |
| Lopinavir/Ritonavir | 80 (34.8) | 30 (37.0) |  |
| Remdesivir | 8 (3.5) | 2 (2.5) |  |
| Tocilizumab | 2 (0.9) | 3 (3.7) |  |
| Anakinra | 23 (10.0) | 8 (9.9) |  |
| LMWH | 176 (76.5) | 76 (93.8) |  |
| Corticosteroids | 33 (14.3) | 17 (21.0) |  |
| Colchicine | 9 (3.9) | 5 (6.2) |  |
| CVVDHF | 0 (0.0) | 4 (4.9) |  |
| **ANC ($\times 10^3$)** |  |  |  |
| Mean ($\pm 95\%$ CI) | 4.6±0.3 | 9.1±1.2 | <0.001 |
| **ALC ($\times 10^3$)** |  |  |  |
| Mean ($\pm 95\%$ CI) | 1.4±0.1 | 1.3±0.2 | 0.177# |
| **NLR** |  |  |  |
| Mean ($\pm 95\%$ CI) | 3.7±0.3 | 11.3±2.2 | <0.001 |
| **CRP (mg/dl)** |  |  |  |
| Mean ($\pm 95\%$ CI) | 8.6±7.9 | 15.8±3.7 | 0.306# |
| **LDH (U/L)** |  |  |  |
| Mean ($\pm 95\%$ CI) | 326±16 | 700±286 | 0.007 |
| **Ferritin (μg/L)** |  |  |  |
| Mean ($\pm 95\%$ CI) | 602±123 | 1213±277 | <0.001 |
| **D-dimers (ng/ml)** |  |  |  |
| Mean ($\pm 95\%$ CI) | 731±131 | 2254±716 | <0.001 |
| **Activin-A** |  |  |  |
| Mean ($\pm 95\%$ CI) | 491±45 | 876±199 | <0.001 |
| **Activin-B** |  |  |  |
| Mean ($\pm 95\%$ CI) | 156±15 | 517±162 | <0.001 |
| **Follistatin** |  |  |  |
| Mean ($\pm 95\%$ CI) | 4127±396 | 12303±3388 | <0.001 |

$\#P<0.001$ using Mann-Whitney test; CVVDHF: continuous venovenous hemodiafiltration; Corticosteroids: dexamethasone or equivalent doses of alternative glucocorticoids; LMWH: low molecular weight heparin; ANC: absolute neutrophil count; ALC: absolute lymphocyte count; NLR: Neutrophil/Lymphocyte Ratio

**Supplementary Table 3.** General linear model for detection of potent confounders.

|  | **F** | **Observed Power** | **P** |
| --- | --- | --- | --- |
| **Multiplicity of samples per patient** |  |  |  |
| Activin-A | 0.858 | 0.152 | 0.355 |
| Activin-B | 1.047 | 0.175 | 0.307 |
| Follistatin | 1.543 | 0.114 | 0.462 |
| **Sampling period (days 1-7/8-14/15+)** |  |  |  |
| Activin-A | 2.188 | 0.445 | 0.114 |
| Activin-B | 1.433 | 0.306 | 0.240 |
| Follistatin | 0.373 | 0.110 | 0.689 |
| **Reference hospital** |  |  |  |
| Activin-A | 1.125 | 0.185 | 0.290 |
| Activin-B | 0.527 | 0.112 | 0.468 |
| Follistatin | 0.437 | 0.101 | 0.509 |

**Supplementary Table 4.** Outcome predictive model as derived using Optimal Scaling procedure along with ridge regression regularization (leading to FACT-CLINYCoD score).

| **FACT-CLINyCoD** | **Cutoff<sup>†</sup>** | **Beta** | **SE estimate<sup>‡</sup>** | **F** | **p** | **Tolerance** |
| --- | --- | --- | --- | --- | --- | --- |
| Activin-A (pg/ml) | 591 | 0.046 | 0.019 | 5.770 | 0.017 | 0.844 |
| Activin-B (pg/ml) | 249 | 0.079 | 0.021 | 13.731 | <0.001 | 0.677 |
| Follistatin (pg/ml) | 6235 | 0.158 | 0.023 | 48.512 | <0.001 | 0.664 |
| Age (years) | 61 | 0.114 | 0.017 | 43.692 | <0.001 | 0.702 |
| NLR | 5.6 | 0.125 | 0.022 | 32.816 | <0.001 | 0.711 |
| CRP (mg/dl) | 10.3 | 0.080 | 0.021 | 14.670 | <0.001 | 0.841 |
| LDH (U/L) | 427 | 0.070 | 0.021 | 11.304 | 0.001 | 0.774 |
| D-dimers (ng/ml) | 1097 | 0.055 | 0.021 | 6.951 | 0.009 | 0.739 |
| ICU admission | - | 0.192 | 0.023 | 71.618 | <0.001 | 0.602 |
| Comorbidities (n) | 1 | 0.049 | 0.016 | 9.580 | 0.002 | 0.710 |

<sup>†</sup> After discretization; <sup>‡</sup> (1000 x bootstrap); ICU: Intensive care Unit

**Supplementary Table 5.** Binary regression for quantification of outcome probability based on the 10-parameter predictive model derived from optimal scaling.

|  | B | SE | Wald | df | p | Exp(B) | -95% CI | +95% CI |
| --- | --- | --- | --- | --- | --- | --- | --- | --- |
| **FACT-CLINyCoDscore** |  |  |  |  |  |  |  |  |
| Points | 1.330 | 0.178 | 56.011 | 1 | <0.001 | 3.781 | 2.669 | 5.357 |
| Constant | -6.198 | 0.766 | 65.455 | 1 | <0.001 | 0.002 |  |  |

**Supplementary Table 6.** Optimal survival using FACT-CLINYCoDscore cut-offs in standard of care (SOC) treatment options.

|  | Responders (%) | Cut-off (stochastic) ‡ |
| --- | --- | --- |
| **LMWH** | 319/437 (73.0) |  |
|  | 206/208 (99.0) | ≤2 |
|  | 113/229 (50.7) | ≥3 |
| **Corticosteroids** | 99/155 (63.9) |  |
|  | 55/56 (98.2) | ≤2 |
|  | 44/99 (44.4) | ≥3 |
| **Remdesivir** | 8/14 (57.1) |  |
|  | 8/8 (100.0) | ND |
|  | 0/6 (0.0) | ND |

ND: Not determined; ‡ Based on ridge regression (all P-values<0.001); CVVDHF: continuous venovenous hemodiafiltration; Corticosteroids: dexamethasone or equivalent doses of alternative glucocorticoids; LMWH: low molecular weight heparin

## SUPPLEMENTARY FIGURES

**Supplementary Figure 1.**
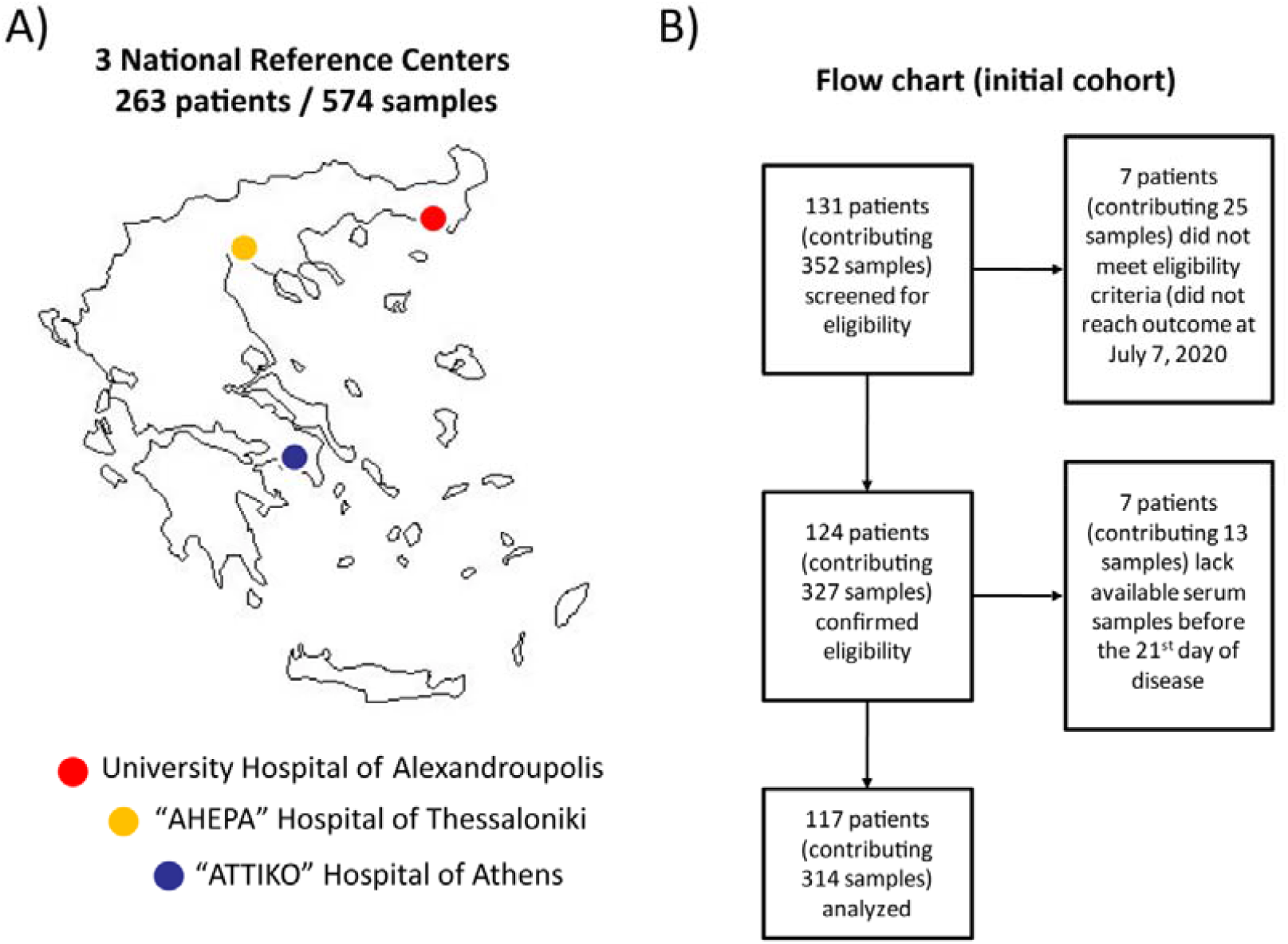
Geographic location of the three reference centers involved in the study (A) and flow chart of patients included in the initial cohort (B).

**Supplementary Figure 2.**
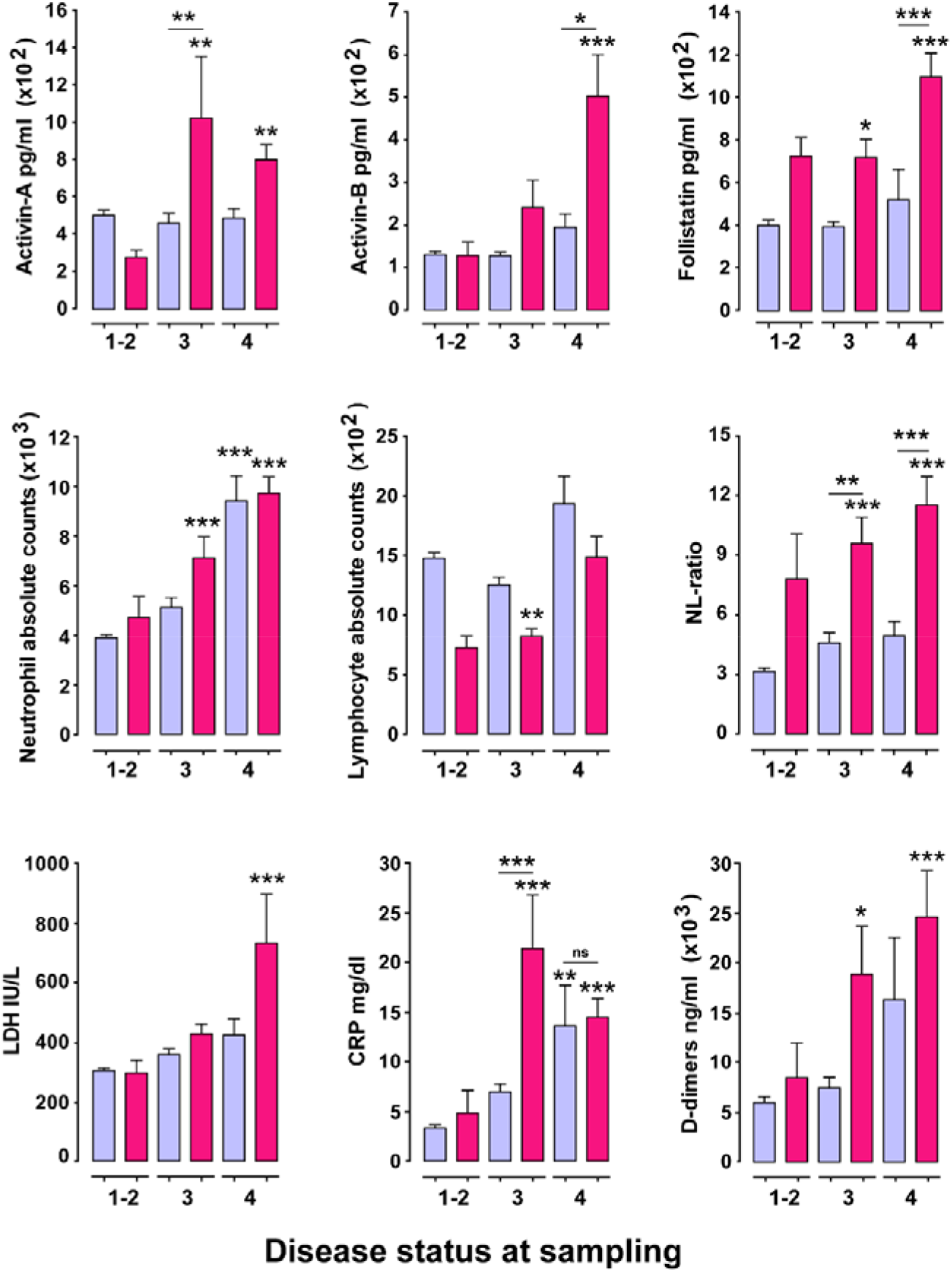
Levels of Activin/Follistatin-axis components are selectively upregulated in non-survivor severe or critically-ill COVID-19 patients. Levels of Activin/Follistatin-axis components in serum and levels of other key inflammatory parameters in the blood of COVID-19 patients grouped according to disease-status (DS) at sampling. Light blue bars represent samples collected from patients that survived COVID-19 and magenta bars represent samples obtained from patients that eventually succumbed to the disease. Data are expressed as mean±SEM analyzed using one-way analysis of variance with Tuckey’s post-hoc test. Asterisks represent comparison of the indicated group with the survivors of DS1-2. Asterisks above a horizontal line represent comparison between the groups under the line segment (*P<0.05, **P< 0.01 and ***P < 0.001).

**Supplementary Figure 3:**
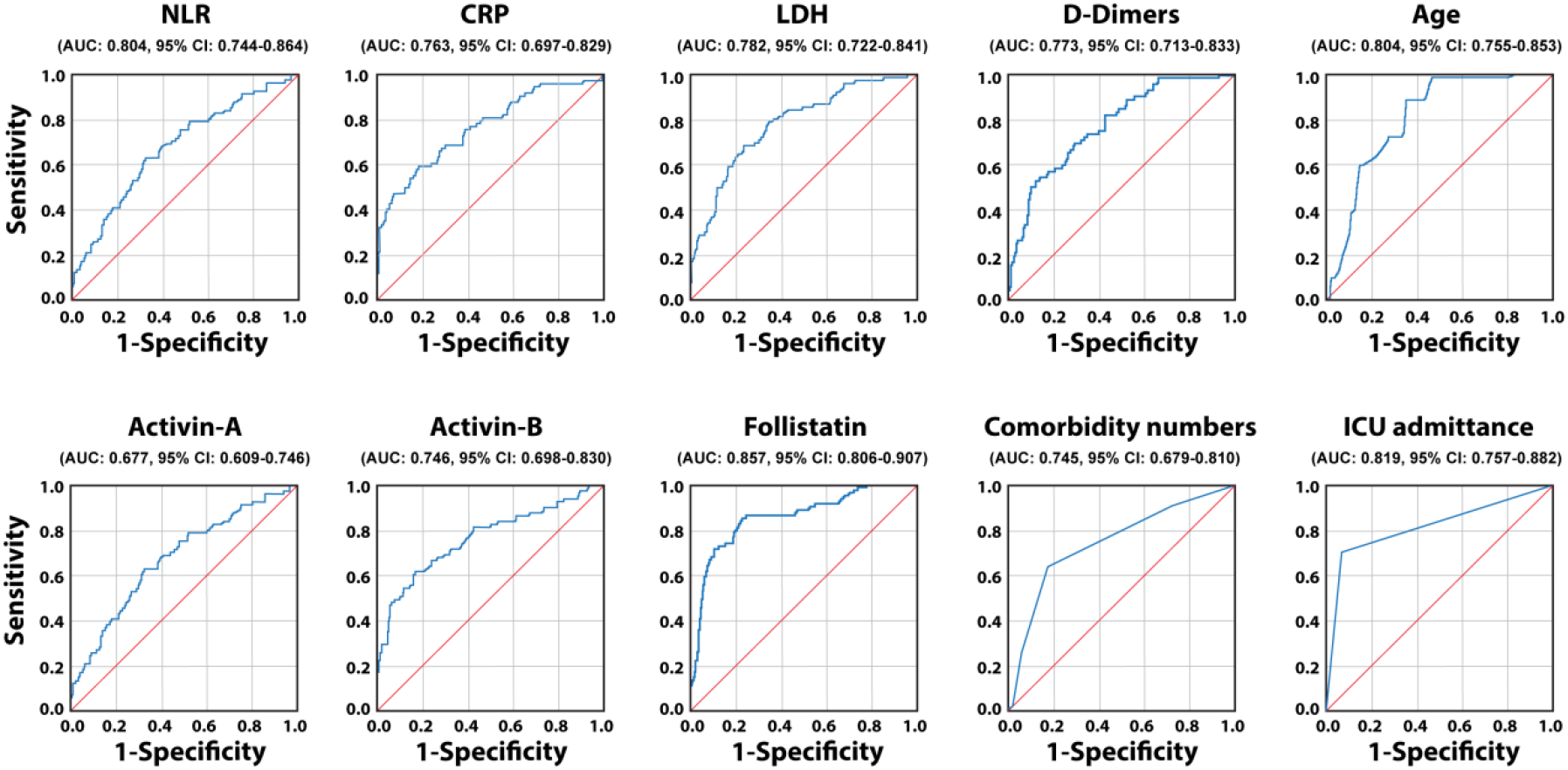
Performance of the indicated single parameters in distinguishing survivor from non-survivor COVID-19 patients. ROC analysis for parameters used in both binary regression and Optimal-Scaling procedures.

**Supplementary Figure 4:**
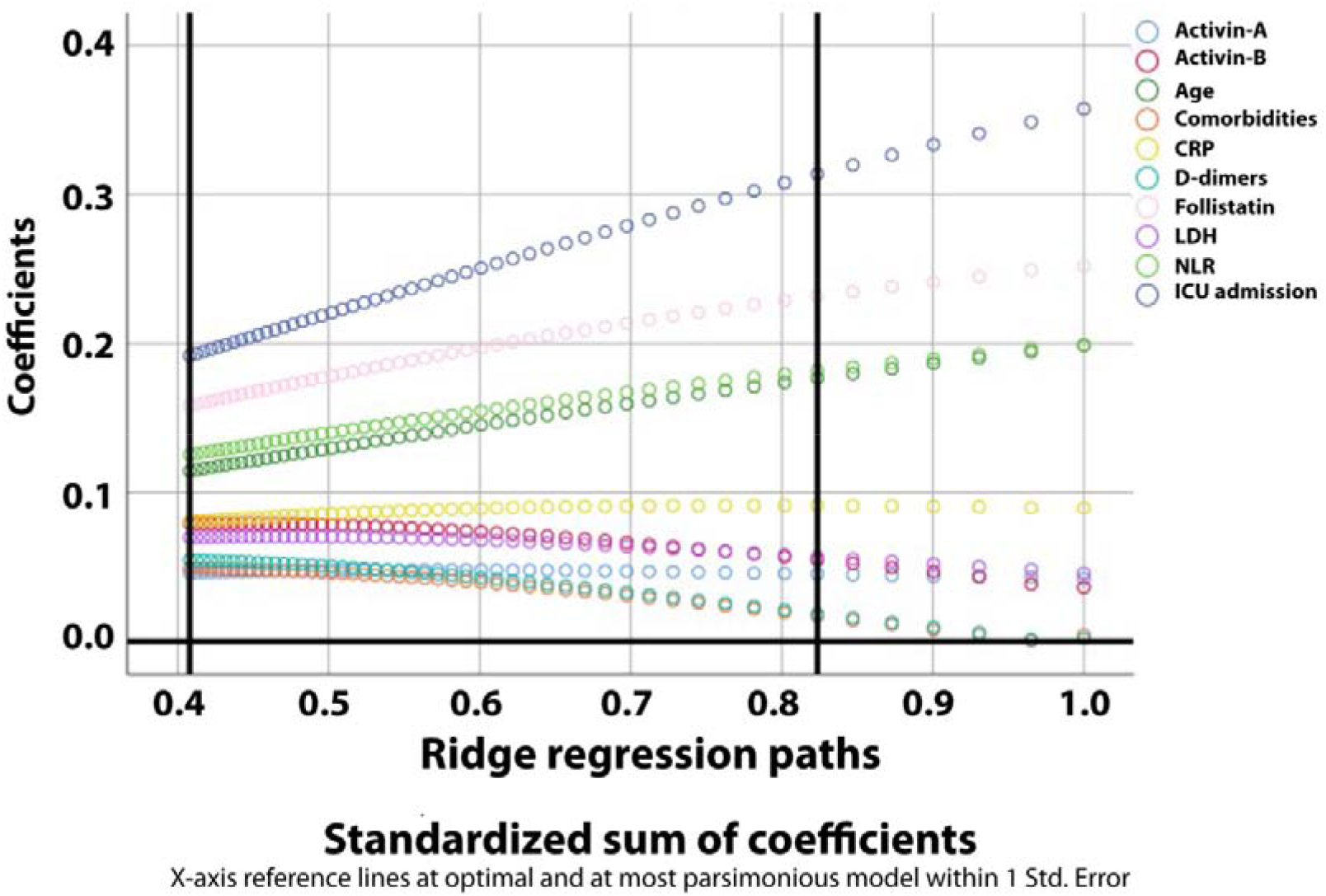
Ridge regression paths for model leading to FACT-CLINYCoD score.

**Supplementary Figure 5:**
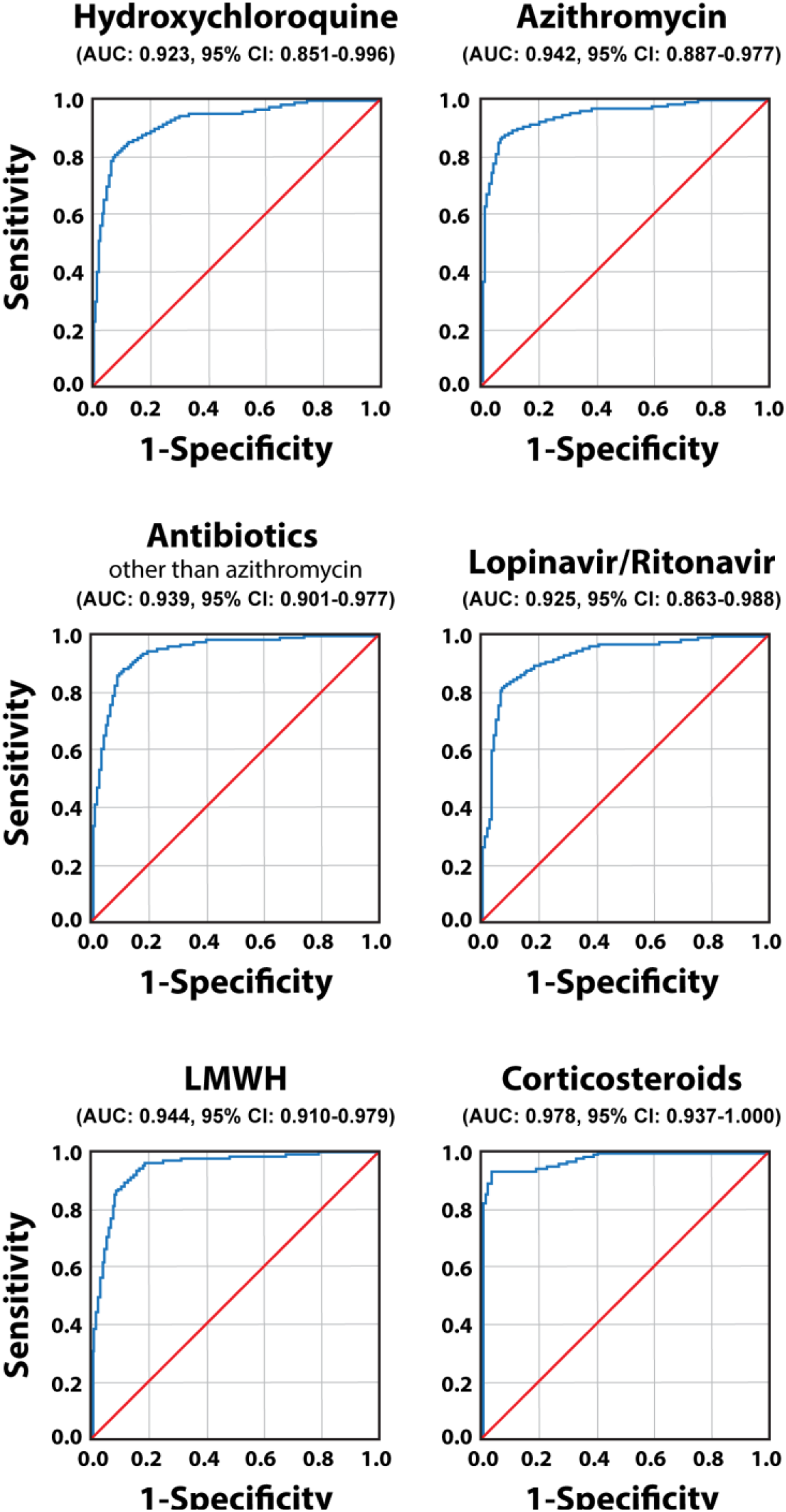
The performance of the FACT-CLINYCoD score in predicting favorable response of COVID-19 patients to the indicated treatments. LMWH: low molecular weight heparin.

